# Seven Decades of Chemotherapy Clinical Trials: A Pan-Cancer Social Network Analysis

**DOI:** 10.1101/19010603

**Authors:** Xuanyi Li, Elizabeth A. Sigworth, Adrianne H. Wu, Jess Behrens, Shervin A. Etemad, Seema Nagpal, Ronald S. Go, Kristin Wuichet, Eddy J. Chen, Samuel M. Rubinstein, Neeta K. Venepalli, Benjamin F. Tillman, Andrew J. Cowan, Martin W. Schoen, Andrew Malty, John P. Greer, Hermina D. Fernandes, Ari Seifter, Qingxia Chen, Rozina A. Chowdhery, Sanjay R. Mohan, Summer B. Dewdney, Travis Osterman, Edward P. Ambinder, Elizabeth I. Buchbinder, Candice Schwartz, Ivy Abraham, Matthew J. Rioth, Naina Singh, Sanjai Sharma, Michael Gibson, Peter C. Yang, Jeremy L. Warner

**Author notes:** Corresponding author: 2220 Pierce Ave, PRB 777, Nashville, TN 37232. Ph. contributed equally (Xuanyi Li, Elizabeth A. Sigworth, Adrianne H. Wu, Peter C. Yang, Jeremy L. Warner).

## Abstract

**Background:** Clinical trials establish the standard of care for cancer and other diseases. While social network analysis has been applied to basic sciences, the social component of clinical trial research is not well characterized. We examined the social network of cancer clinical trialists and its dynamic development over more than 70 years, including the roles of subspecialization and gender in relation to traditional and network-based metrics of productivity.

**Methods:** We conducted a social network analysis of authors publishing chemotherapy-based prospective trials from 1946-2018, based on the curated knowledge base HemOnc.org, examining: 1) network density; 2) modularity; 3) assortativity; 4) betweenness centrality; 5) PageRank; and 6) the proportion of co-authors sharing the same primary cancer subspecialty designation. Individual author impact and productive period were analyzed as a function of gender and subspecialty.

**Findings:** From 1946-2018, the network grew to 29,197 authors and 697,084 co-authors. While 99.4% of authors were directly or indirectly connected as of 2018, the network had very few connections and was very siloed by cancer subspecialty. Small numbers of individuals were highly connected and had disproportionate impact (scale-free effects). Women were under-represented and likelier to have lower impact, shorter productive periods (P<0.001 for both comparisons), less centrality, and a greater proportion of co-authors in their same subspecialty. The past 30 years were characterized by a trend towards increased authorship by women, with new author parity anticipated in 2032. However, women remain a distinct minority of first/last authors, with parity not anticipated for 50+ years.

**Interpretation:** The network of cancer clinical trialists is best characterized as a strategic or “mixed-motive” network, with cooperative and competitive elements influencing its appearance.

Network effects e.g., low centrality, which may limit access to high-profile individuals, likely contribute to ongoing disparities.

**Funding:** Vanderbilt Initiative for Interdisciplinary Research; National Institutes of Health; National Science Foundation

**Research in context:** *Evidence before this study:* We reviewed the literature on social networks from the 1800’s to 2018. Additionally, MEDLINE was searched for (“Social Networking”[Mesh] OR “Social Network Analysis”) AND (“Clinical Trials as Topic”[Mesh] OR “Hematology”[Mesh] OR “Medical Oncology”[Mesh]) without date restriction. The MEDLINE search yielded 43 results, of which 8 were relevant; none considered gender nor temporality in their analyses. To our knowledge, there has not been any similar study of the dynamic social network of clinical trialists from the inception of the fields of medical oncology and hematology to the present.

*Added value of this study:* This is the first dynamic social network analysis of cancer clinical trialists. We found that the network was sparse and siloed with a small number of authors having disproportionate impact and influence as measured by network metrics such as PageRank; these metrics have become more disproportionate over time. Women were under-represented and likelier to have lower impact, shorter productive periods, less network centrality, and a greater proportion of co-authors in their same cancer subspecialty.

*Implications of all the available evidence:* While gender disparities have been demonstrated in many fields including hematology/oncology, our analysis is the first to show that network factors themselves are significantly implicated in gender disparity. The increasing coalescence of the network by traditional cancer type and around a small number of high-impact individuals implies challenges when the field pivots from traditionally disease-oriented subspecialties to a precision oncology paradigm. New mechanisms are needed to ensure diversity of clinical trialists.

## Background and Motivation

The modern era of chemotherapy began in 1946, with publications describing therapeutic uses of nitrogen mustard.^1,2^ Over the next 70 years, the repertoire of cancer treatment has expanded at an ever-increasing pace. Chemotherapies have a notably low therapeutic index, making them the most dangerous interventions commonly used in biomedicine.^3^ Consequently, a complex international clinical trial apparatus emerged in the 1970s and prospective clinical trials remain the gold standard by which standard of care treatments are established.^4,5^ Discoveries made by successive generations have led to overall improvement in the prognosis of most cancers.^6^

While social network analysis has been used in scientific settings,^7,8^ the social component of clinical trial research is not well characterized. How social dynamics have influenced progress over time, as cancer care has become increasingly subspecialized, remains to be understood. Little is known about how social factors have shaped the field, and how social network characteristics may reveal patterns of inclusiveness, exclusivity, and disparity. We hypothesized that the social network of cancer clinical trialists would be a “strategic” aka “mixed motive” network, where collaborative and competitive pressures are counterbalanced; this type of social network may be especially prone to preferential attachment and social disparities.^9,10^

Here, we analyzed the individual impact and collaborative relationships of cancer clinical trialists, using co-authorship to define social networks. Our primary objective was to model the social network and its dynamic development over time; secondary objectives were to examine the roles of subspecialization and gender in relation to metrics of productivity.

## Methods

All prospective trials of systemic antineoplastics published between 1946-2018 and referenced on HemOnc.org were considered for inclusion. HemOnc.org is the largest collaborative wiki of chemotherapy drugs and regimens and has a formal curation process (**eMethods**).^11^ For each included publication, author names were extracted, disambiguated, and mapped to gender (**eMethods** and **Supplemental Files 1 & 2**). Every author was assigned an impact score, using an algorithm based on four aspects of their publications: author role (first/last versus middle author); citations; trial design (randomized versus non-randomized); and whether the publication was primary or an update (**eMethods**).

Each publication was assigned to disease-specific subspecialties based on the cancer(s) studied (**eTable 1**), and authors were assigned a primary subspecialty based on impact (**eMethods**).

Author longevity was calculated as the interval between their first and final publication in the database. Given that preparing and publishing results of clinical trials can take substantial time, for authors with first publication prior to 2016 and final in 2016 or 2017, final was adjusted to 2018.

### *Social network metrics* (eGlossary)

A dynamic social network was created with nodes representing authors and links representing co-authorship. The dynamic social network was discretized by year and the authors, scores, and links were cumulative (e.g., the 20^th^ network was cumulative from 1946-1965). Therefore, once an author is added to the network, they remain in the network, with their impact score cumulatively increasing as they publish and remaining constant if publication activity ceases. The following temporal metrics were calculated: 1) network density; 2) modularity^12^ by subspecialty; 3) assortativity^13^ by subspecialty; 4) betweenness centrality^14^; 5) PageRank^15^; and 6) proportion of co-authors sharing the same primary subspecialty designation (hereafter – homophily). All metrics except homophily incorporated the weighted co-authorship score, which takes into account each co-author’s impact modified by the number of authors of an individual publication (see **eMethods**). In order to visualize the final cumulative network, layout was determined using the distributed recursive graph algorithm.^16^ Nodes were sized by author impact score rank and colored by primary subspecialty designation. Edge width was determined by the weighted co-authorship score.

### Statistical analysis

Non-independent network metrics including growth, density, assortativity, modularity, and PageRank are reported descriptively with medians and interquartile ranges (IQR). Gender proportion over time was fit with locally estimated scatterplot smoothing (LOESS) regression using default settings of degree=2 with smoothing parameter/span α=0.75.^17^ For the final cumulative network, the independent variables author impact score and longevity were compared 1) between genders and 2) by whether the author changed subspecialties over time; only those authors with longevity ≥1 year were included in the second comparison. These comparisons were made with the two-sided Wilcoxon rank sum test; P value <0.05 was considered statistically significant.

### Sensitivity analysis

To determine whether the scoring algorithm was robust to modifications, we conducted a sensitivity analysis where the author role and trial design coefficients were varied by +/-67% and +/-50%, respectively. Normalized density distributions for the final cumulative network under each permutation were calculated, and temporal assortativity and modularity were compared to baseline with Pearson’s correlation coefficient.

## Results

### Baseline characteristics

N=5,599 of 6,301 reviewed publications with an aggregate of n=29,197 authors met inclusion criteria (CONSORT **eFigure 1**). Cumulatively, most authors (78%) participated in publication of a randomized trial, with 52.5% participating in the publication of a “positive” trial (**eTable 2**). The median number of authors per publication increased from six in 1946 to 20 (IQR 16-25) in 2018 (**eFigure 2**).

The final network is very sparse (0.16% of possible links are present); nevertheless, n=29,029 (99.4%) authors are in a single connected component; the next-largest component comprises 14 authors.

### Network dynamics

Authorship and co-authorship have grown by over 200,000%: in 1946 there were 12 authors & 30 co-authors; by 2018, there were cumulatively 29,197 authors & 697,084 co-authors (**Figure 1A**). Median longevity is <1 year at all times, although the number of authors with multiple years in the field grows substantially over time (**eFigure 3**). A small number of individuals maintained the highest impact over time – nearly 20 years each in the case of chemotherapy pioneers Sidney Farber and James F. Holland (**eFigure 4**). In any given year, most authors had a betweenness centrality of <1% of the maximum; conversely, a very small number of authors had an exceptionally high score, with 1% of authors accounting for 100% of the total in recent years (**eFigure 5**). Accordingly, an increasingly smaller proportion of authors were both very highly connected and highly impactful; e.g., in 1970, the 10% highest-impact authors (N=20) account for 21.4% of links and 54.9% of impact; in 2018, the same proportion (N=2,920) account for 37.1% of links and 62.3% of impact. First/last authorship has also become concentrated; in 2018 publications, 10% of authors had at least one such role, whereas prior to 1980 it was on average >25% (**eFigure 6**).

**Figure 1.**
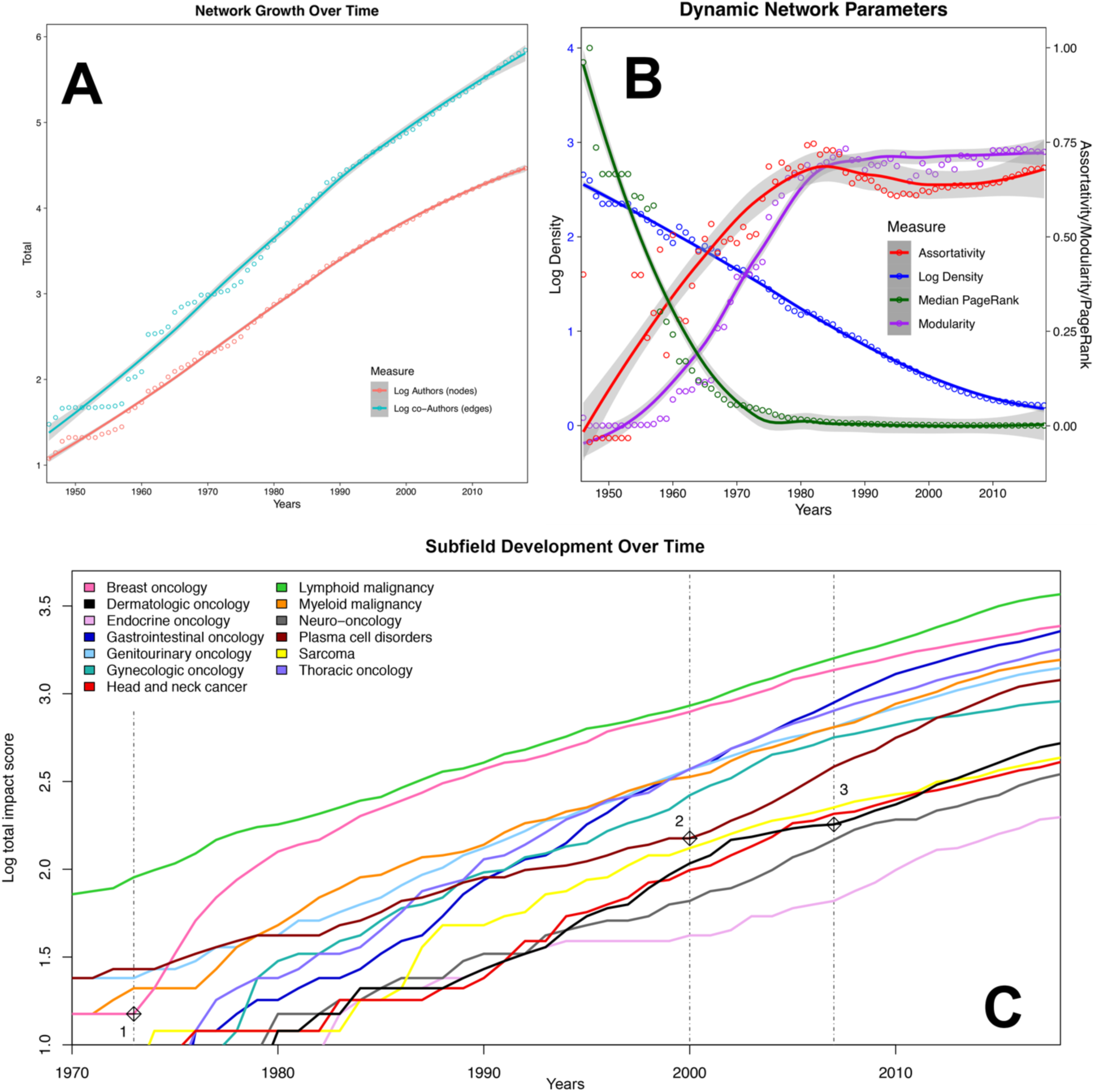
Network characteristics. A: Cumulative growth in authorship and co-authorship over time have both been nearly log-linear; B: Network density decreases asymptotically from 45.5% in 1946 to 0.16% in 2018; modularity follows a sigmoid pattern with a period of linear increase between 1960-80 followed by a plateau at high modularity; assortativity rapidly increases in early decades; median normalized PageRank decreases to a low plateau from the 1970s onward; C: Subspecialties develop at different but broadly parallel rates, with seminal events apparently preceding accelerations of individual subspecialties, e.g.,: 1) in the four years after 1973, combination therapy (AC^18^), adjuvant therapy^19^, and tamoxifen^20^ were introduced in breast cancer; 2) thalidomide^21^ and bortezomib^22^ were reported to be efficacious for multiple myeloma; and 3) immunotherapy (ipilimumab^23,24^) was introduced in the treatment of melanoma.

The structure of the network changes considerably over time, from relatively dense and connected to sparse and modular (**Figure 1B**). Each of thirteen cancer subspecialties develops at different rates, with clear influence of seminal events in several subspecialties, e.g., the introduction of adjuvant therapy and tamoxifen for breast cancer, completely new classes of drugs for plasma cell disorders, and immunotherapy for melanoma (**Figure 1C**).^18–24^

### Network visualization and cumulative metrics

The final cumulative network visualization is shown in **Figure 2**. The impact score of authors is unevenly distributed, median 0.0532 (range 0-14.31); however, the log-transformed impact scores approximate a normal distribution (**eFigure 7**). Authors with longevity ≥1 year who changed primary subspecialty at least once (n=2,330) had nearly twice the median impact and longevity of those who remained in one subspecialty (n=10,276), 0.25 (IQR 0.11-0.6) versus 0.14 (IQR 0.07-0.35) and 13 years (IQR 6-19) versus 7 years (IQR 3-12), respectively (P<0.001 for both comparisons).

**Figure 2.**
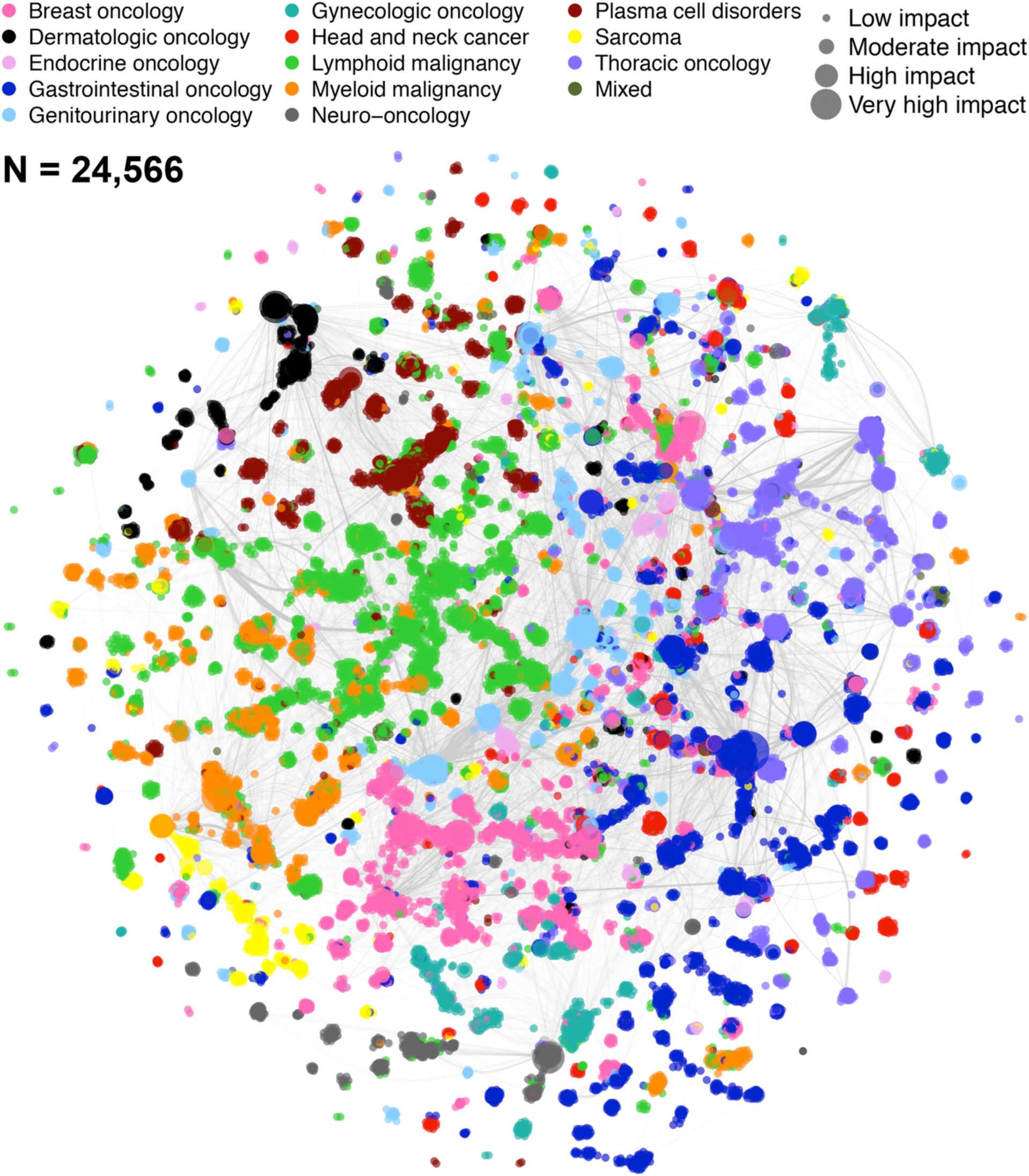
Final cumulative network visualization. The social network graph represents the cumulative field of cancer research as of December 31, 2018, with all included published works since 1946 contributing to authorship and co-authorship weights. Only authors assigned to a subspecialty are visualized; these account for 84% of all authors in the database.

Cumulatively, subspecialized authors with calculable homophily (n=24,560) have a median proportion of co-authors sharing the same subspecialty of 88% (IQR 76-95%); 945,167 (71.4%) of these authors’ outlinks are within-subspecialty. This is reflected by a high assortativity by subspecialty since the mid-1960s (**Figure 1B**).

### Gender disparities

The proportion of woman authors remained at a nearly steady state of 15% until 1980, when it began to gradually increase to a recent high of ∼40%; over the same time period, the proportion of woman first/last authors rose much more slowly (**Figure 3A**). Cumulatively, n=17,187 men had a statistically significantly higher median impact than n=8,511 women, 0.075 (IQR 0.032-0.22) versus 0.051 (IQR 0.022-0.133), P<0.001; statistically significantly longer median longevity, 1 year (IQR 0-8) versus 0 years (IQR 0-5), P<0.001; and higher median PageRank, 1.75 x 10^−5^ (IQR 1.01 x 10^−5^-3.68 x 10^−5^) versus 1.34 x 10^−5^ (IQR 8.74 x 10^−6^-2.51 x 10^−5^). For the n=15,229 men and n=7,245 women with a calculable homophily, men had a lower median proportion of co-authors within the same primary subspecialty, 0.88 (IQR 0.75-0.95) versus 0.89 (IQR 0.78-0.95). Scatterplots of longevity versus author impact score and PageRank versus homophily are shown in **Figures 3B & 3C** for the final cumulative network; prior years are shown in **eFigures 8 & 9**.

**Figure 3.**
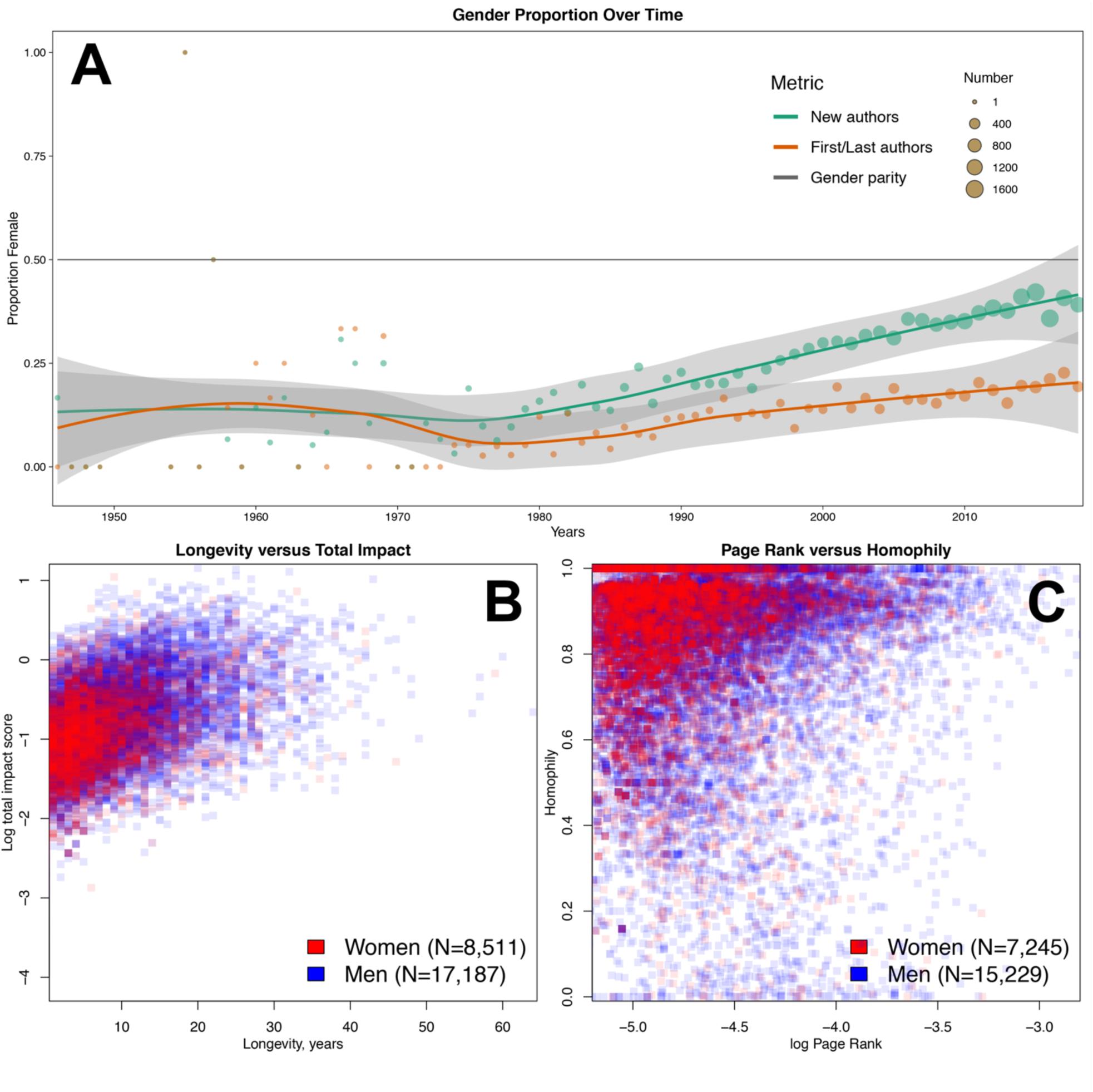
Gender disparities in the network. A: The network is overwhelmingly dominated by men until 1980, when a trend towards increasing authorship by women begins to be seen; however, representation by women in first/last authorship remains low; gray shaded lines are 95% confidence intervals of the LOESS curves; B: Men tend on average to have a longer productive period and to achieve a higher author impact score than women (P<0.001 for both comparisons); C: Men tend on average to be more central and have more collaborations outside of their subspecialty. Note that the homophily calculation requires a subspecialty assignment, which explains the slightly lower numbers in 3C as compared to 3B.

### Sensitivity analysis

Normalized score distributions did not change significantly, although modulation of the trial design coefficient led to a bimodal peak (**eFigure 10**). Correlation of assortativity and modularity was high, ranging from 0.815-0.999 for the former and 0.981-0.999 for the latter (**eTable 3**; **eFigure 11**).

## Discussion

The remarkable gains in the fields of hematology and oncology can be ascribed to the tireless work of numerous trialists and the generosity of countless patient participants. As a result, systemic antineoplastics now stand beside surgery and radiotherapy as a pillar of cancer care. Our analysis of clinical trialists as a social network reveals a mixed-motive network that differs substantially from “collegial” and “friend-based” commercial online social networks. While clinical trials are conducted towards a collaborative goal – improved outcomes for all cancer patients – there are significant competitive pressures. Examples of these pressures include resource limitations (e.g., funding and patients available for accrual), the tension between prioritization of cooperative group versus industry-funded trials, personal motivations such as academic promotion or leadership opportunities, and institutional reputation.

The emergence of formal and informal leaders in scientific networks has been shown to facilitate research as well as create silos.^25^ As **Figure 2** shows, there is a strong tendency for siloing based on subspecialty in the complete network, although some subspecialties (e.g., lymphoid and myeloid malignancies) have many more interconnections than others (e.g., sarcoma and neuro-oncology). Many of these silos appear to be organized around an individual or group of individuals who have high impact and centrality. As an organizational principle, these individuals appear to rarely be in direct competition, but their presence is a clear indicator of scale-free phenomena within the network. The facts that betweenness follows a power law cumulative distribution bolsters this theory. Scale-free phenomena, which are defined by a power law distribution of connectedness, are very common in strategic networks, especially when they become increasingly sparse, as this network does.^26^ The two related theories for this network behavior are preferential attachment and fitness. The former observes that those with impact tend to attract more impact; the latter postulates that such gains for the “fittest” come at the expense of the “less fit.”^27^ Seminal events (**Figure 1C**) are likely a driver of preferential attachment,^28^ and may partially explain why some authors change their primary subspecialty at least once over time (e.g., through a “bandwagon” effect driven by the diffusion of ideas^29^). Given that these authors were observed to have nearly twice the impact and longevity of their single subspecialty peers, this dynamic will be a focus of future study, including calculation of the Q factor described by Sinatra et al.^30^

In the analysis of network dynamics (**Figure 1B**), the field as a whole appears to emerge in the 1970s, which is also when formal recognition of medical oncology and hematology through board certification occurred. Measurements of field maturity are by their nature subjective, but the pessimism^31^ of the late 1960s was captured by Sidney Farber: “…the anticancer chemicals, hormones, and the antibiotics…marked the beginning of the era of chemotherapy of cancer, which may be described after 20 years as disappointing because progress has not been more rapid…”.^32^ These concerns led directly to the US National Cancer Act of 1971, which was followed by the leveling of modularity at a very high level from 1976 onwards, suggesting that the subspecialties generated in the 1970s have remained stable. The assortativity by subspecialty has increased as well, with recent levels approximately twice those seen in a co-authorship network of physicists.^33^ While median PageRank has decreased markedly, indicating decreasing influence for the average author, the distribution in 2018 is broadly right-skewed (**eFigure 12**). These findings reveal a high level of exclusivity, which has been increasing over time, suggesting that it is becoming increasingly difficult to join the top echelon of the network. This has major implications for junior investigators’ mobility, and potentially the continued health of the network as a whole.

While there is much to be applauded in the continued success of translating research findings into the clinic, we observed clear gender disparities within the cancer clinical trialist network. Women have a statistically significantly lower final impact score, shorter productive period, less centrality, and less collaboration with those outside of their primary subspecialty. These findings are consistent with and build upon previous literature on the challenges facing women academics.^34,35^ Others studies investigating the basis for such a gender gap have identified several layers of barriers to the advancement of women in academic medicine. These include sexism in the academic environment, lack of mentorship, and inequity with regards to resource allocation, salary, space, and positions of influence.^36,37^ Our study suggests that additional network factors such as relatively low centrality, which indicates a lack of access to other individuals of influence, and high homophily, which indicates a lack of access to fresh ideas and perspectives, also perpetuate the gender gap – corroborating recent findings from graduate school social networks.^38^

It is somewhat encouraging that there has been a steady increase in the proportion of authorship by women since 1980 (**Figure 3A**). This increase is observed approximately 10 years after the passage of Title IX of the US Civil Rights Act in 1972. Given that the majority of authors in this network are clinicians, a partial explanation is that US-based women began to attend previously all-male medical schools in the early 1970s, completed their training, and began to contribute to the network as authors approximately 10 years later. If the nearly linear trend continues, we predict that gender parity for new authors entering the network will be reached by the year 2032, 26 years after US medical school enrollment approached parity.^39^ However, the proportion of woman first/last authors is growing much more slowly, and parity may not be reached for 50+ years, if at all. Given that senior authorship is a traditional metric of scholarly productivity, it may be particularly difficult for clinical trialists who are women to obtain promotion under the current paradigm. One possible solution is to increase the role of joint senior authorship, which remains vanishingly rare in the clinical trials domain (Furman et al. 2014^40^ is one of very few examples that we are aware of) – although this is predicated on the acceptance of these roles by advancement and promotion committees. The field itself may also suffer from slow entry of new talent and a lack of broad perspectives.

While the gender mapping algorithm and manual lookups are imperfect, our approach is consistent with prior work in this area.^34,41^ Unisex names posed a particular challenge.^42^ It should be noted that we could not account for all situations where an author changed their name (e.g., a person assumed their spouse’s surname); this could have led to overestimation of representation by women and underestimation of impact, since this practice is more common with women. It is also possible that an individual’s gender identity does not match the gender assignment of their given name. Future work will include further analysis of gender disparities, factoring in institutional affiliation and highest degree(s) obtained, which are both likely to have significant influence on publication and senior authorship.^43,44^

There are several additional limitations to this work, starting with the fact that co-authorship is but one way to measure social network interactions and this study reports results from published trials, which induces publication bias. Although HemOnc.org aims to be the most comprehensive resource of its kind, non-randomized and randomized phase II trials are intentionally underrepresented, given that findings at this stage of investigation infrequently translate to practice-changing results (e.g., approximately 70% of oncology drugs fail during phase II).^45–47^ The effect of any biases introduced by this underrepresentation is unclear, given the confounding influence of publication bias, which may itself be subject to gender disparity.^48^ Some older literature which no longer has practice-changing implications may have been overlooked.

During name disambiguation, some names could not be resolved, primarily because neither MEDLINE nor the primary journal site contained full names. This effect is non-random, since certain journals do not publish full names. The choice of coefficients and their relative weights was based on clinical intuition and consensus; given that the “worth” of metrics such as first/last authorship is fundamentally qualitative, there must be some degree of subjectivity when formulating a quantitative algorithm. While the sensitivity analysis demonstrated that neither normalized author impact score distribution, assortativity, nor modularity are majorly changed by variation in the trial design and author role coefficients, it remains possible that other combinations of coefficients and relative weightings could lead to different results. Furthermore, our impact algorithm weighs heavily on first and last authorship, but the definition of senior authorship has changed over time. For example, in the 1946 article by Goodman et al.,^2^ the authors were listed in decreasing order of seniority (personal communication). Finally, the majority of authors in this database publish extensively and their impact as measured here should not be misconstrued to reflect their contributions to the cancer field more broadly.

In conclusion, we have described the first and most comprehensive social network analysis of the clinical trialists involved in chemotherapy trials. We found emergent properties of a strategic network, as well as clear indications of gender disparities, albeit with some improvement in representation in recent decades. The network has been highly modular and assortative for the past 40 years, with little collaboration across most subspecialties. As hematology and oncology pivot from an anatomy-based to a precision medicine paradigm, it remains to be seen how the network will re-organize so that the incredible progress seen to date can continue.

## Data Availability

Disambiguated author names and names mapped to gender are available as supplemental files. Algorithms are available upon request.

## Author contributions

A.H.W., P.C.Y., and J.L.W. conceived and designed the study. A.H.W. and J.L.W. developed and implemented the name disambiguation algorithms. X.L. developed and implemented the automated gender mapping algorithms. A.M. developed and implemented the citation count algorithm. E.A.S., J.B., Q.C., and J.L.W. contributed to the network analysis. X.L., A.H.W., S.A.E., S.N., R.S.G, K.W., E.J.C., S.M.R., N.K.V., B.F.T., and A.J.C. contributed to manual gender mapping. S.N, R.S.G., E.J.C., S.M.R., N.K.V, B.F.T., A.J.C., M.W.S., J.P.G., H.D.F., A.S., R.A.C., S.R.M., S.B.D., T.J.O., E.P.A., E.I.B., C.S., I.A., M.J.R., N.S., S.S., and M.G. contributed clinical content expertise. All authors contributed intellectual content during the drafting and revisioning of the work and approved the final version.

## Non-author contributions

We thank Ja Young Kwon, MD, PhD (Yonsei University College of Medicine) and David Y. Suh, BA (Vanderbilt University School of Medicine) for their assistance with Korean name gender mapping. We also thank Harry Hochheiser, PhD (University of Pittsburgh), Allison H. Schachter, PhD (Vanderbilt University), Philip Walker, MLIS, MSHI (Vanderbilt University), and Yaomin Xu, PhD (Vanderbilt University) for their review and/or suggestions, and all of the current and former HemOnc.org editorial board members for their contributions to the site. No compensation has been received by any of the persons listed here.

## Access to data and data analysis

Jeremy L. Warner had full access to all the data in the study and had final responsibility for the integrity of the data, the accuracy of the data analysis, and the decision to submit for publication. Xuanyi Li (Vanderbilt University), Elizabeth A. Sigworth (Vanderbilt University), Jess Behrens (Vanderbilt University), Andrew Malty (Stanford University), Qingxia Chen (Vanderbilt University), and Jeremy L. Warner (Vanderbilt University) conducted and are responsible for the data analysis.

## Originality of content

All information and materials in the manuscript are original.

